# More than the sum of its parts: disrupted core-periphery of multiplex networks in multiple sclerosis

**DOI:** 10.1101/2022.12.17.22283623

**Authors:** Giuseppe Pontillo, Ferran Prados, Alle Meije Wink, Baris Kanber, Alvino Bisecco, Tommy A. A. Broeders, Arturo Brunetti, Alessandro Cagol, Massimiliano Calabrese, Marco Castellaro, Sirio Cocozza, Elisa Colato, Sara Collorone, Rosa Cortese, Nicola De Stefano, Linda Douw, Christian Enzinger, Massimo Filippi, Michael A. Foster, Antonio Gallo, Gabriel Gonzalez-Escamilla, Cristina Granziera, Sergiu Groppa, Hanne F. Harbo, Einar A. Høgestøl, Sara Llufriu, Luigi Lorenzini, Eloy Martinez-Heras, Silvia Messina, Marcello Moccia, Gro O. Nygaard, Jacqueline Palace, Maria Petracca, Daniela Pinter, Maria A. Rocca, Eva Strijbis, Ahmed Toosy, Paola Valsasina, Hugo Vrenken, Olga Ciccarelli, James H. Cole, Menno M. Schoonheim, Frederik Barkhof, the MAGNIMS study group

**Affiliations:** Queen Square Multiple Sclerosis Centre, Department of Neuroinflammation, UCL Queen Square Institute of Neurology, University College London, London, United Kingdom; MS Center Amsterdam, Radiology and Nuclear Medicine, Vrije Universiteit Amsterdam, Amsterdam Neuroscience, Amsterdam UMC location VUmc, Amsterdam, The Netherlands; Departments of Advanced Biomedical Sciences and Electrical Engineering and Information Technology, University of Naples “Federico II”, Naples, Italy; Centre for Medical Image Computing, Department of Medical Physics and Biomedical Engineering, University College London, London, United Kingdom; E-Health Center, Universitat Oberta de Catalunya, Barcelona, Spain; Department of Advanced Medical and Surgical Sciences, University of Campania “Luigi Vanvitelli”, Naples, Italy; MS Center Amsterdam, Anatomy and Neurosciences, Vrije Universiteit Amsterdam, Amsterdam Neuroscience, Amsterdam UMC location VUmc, Amsterdam, The Netherlands; Translational Imaging in Neurology (ThINK) Basel, Department of Biomedical Engineering, Faculty of Medicine, University Hospital Basel and University of Basel, Basel, Switzerland; Department of Neurology, University Hospital Basel, Switzerland; Research Center for Clinical Neuroimmunology and Neuroscience Basel (RC2NB), University Hospital Basel and University of Basel, Basel, Switzerland; Department of Neurosciences, Biomedicine and Movement Sciences, University of Verona, Verona, Italy; Department of Information Engineering, University of Padova, Padova, Italy; Department of Medicine, Surgery and Neuroscience, University of Siena, Siena, Italy; 14Department of Neurology, Medical University of Graz, Graz, Austria; Movement Disorders, Neurostimulation and Neuroimaging, University Medicine Mainz, Mainz, Germany; Department of Neurology, Oslo University Hospital, Oslo, Norway; Department of Psychology, University of Oslo, Oslo, Norway; Center of Neuroimmunology. Laboratory of Advanced Imaging in Neuroimmunological Diseases; Hospital Clinic Barcelona, Institut d’Investigacions Biomediques August Pi i Sunyer (IDIBAPS) and Universitat de Barcelona. Barcelona, Spain.; Nuffield Department of Clinical Neurosciences, University of Oxford, Oxford, United Kingdom; Department of Neurosciences and Reproductive and Odontostomatological Sciences, University of Naples “Federico II”, Naples, Italy; Neuroimaging Research Unit, Division of Neuroscience, IRCCS San Raffaele Scientific Institute, Milan, Italy; Neurology Unit, IRCCS San Raffaele Scientific Institute, Milan, Italy; Neurorehabilitation Unit, IRCCS San Raffaele Scientific Institute, Milan, Italy; Neurophysiology Service, IRCCS San Raffaele Scientific Institute, Milan, Italy; Vita-Salute San Raffaele University, Milan, Italy; MS Center Amsterdam, Neurology, Vrije Universiteit Amsterdam, Amsterdam Neuroscience, Amsterdam UMC location VUmc, Amsterdam, The Netherlands; Centre for Medical Image Computing, Department of Computer Science, University College London, London, United Kingdom; Dementia Research Centre, UCL Queen Square Institute of Neurology, University College London, London, United Kingdom

**Author notes:** Correspondence to: Giuseppe Pontillo, MD, PhD, Queen Square MS Centre, UCL Institute of Neurology, National Hospital for Neurology & Neurosurgery, Queen Square, London WC1N 3BG, UK. **Abbreviations:** BOLD = blood oxygenation level dependent; BPF = brain parenchymal fraction; CIS = clinically isolated syndrome; CSF = cerebrospinal fluid; dMRI = diffusion MRI; DMT = disease-modifying treatment; EDSS = Expanded Disability Status Scale; FC = functional connectivity; GM = gray matter; HC = healthy controls; MC = morphological covariance; PPMS = primary-progressive multiple sclerosis; RRMS = relapsing-remitting multiple sclerosis; rs-fMRI = resting-state functional MRI; SC = structural connectivity; sMRI = structural MRI; SPMS = secondary-progressive multiple sclerosis; PwMS = people with multiple sclerosis; SDMT = Symbol Digit Modalities Test; TIV = total intracranial volume; WM = white matter.

**Keywords:** multiple sclerosis, MRI, brain connectivity, multilayer networks, core-periphery structure.

## Abstract

Disruptions to brain networks, measured using either structural (sMRI), diffusion (dMRI), or resting-state functional (rs-fMRI) MRI, have been shown in people with multiple sclerosis (PwMS), highlighting the importance of damage to regions in the core of the connectome. Here, using a multilayer network approach, we aimed to integrate these three modalities to portray an enriched representation of the brain’s core-periphery organization and explore its alterations in PwMS.

In this retrospective cross-sectional study, 1048 PwMS (695F, mean±SD age: 43.3±11.4yr), and 436 healthy controls (250F, mean±SD age: 38.3±11.8yr) with complete multimodal brain MRI acquisitions were selected from 13 European centres within the MAGNIMS network. Clinical variables included the Expanded Disability Status Scale (EDSS) and the Symbol Digit Modalities Test (SDMT), measuring physical disability and cognition, respectively. SMRI, dMRI, and rs-fMRI data were parcellated into 100 cortical (Schaefer atlas) and 14 subcortical (FSL-FIRST) regions to obtain networks of morphological covariance, structural connectivity, and functional connectivity, respectively. Following statistical harmonization and preprocessing, connectivity matrices were merged in a multiplex, from which regional coreness, defined as the probability of a node being part of the multiplex core, and coreness disruption index (κ), quantifying the global weakening of the core-periphery structure, were computed.

The associations of regional coreness and κ with disease status (PwMS versus healthy controls), clinical phenotype, and physical (EDSS) and cognitive (SDMT z-scores) disability were tested with permutation testing, one-way ANOVA, and Spearman and Pearson correlation, respectively. We used random forest permutation feature importance to assess the relative weights of κ in the multiplex and single-layer domains, in addition to conventional MRI measures (brain and lesion volumes), for the prediction of disease status, level of physical disability (EDSS≥4 vs EDSS<4), and cognitive impairment (SDMT z-score<-1.5).

PwMS showed widespread deviations in regional coreness compared to healthy controls, with a prominent decrease in the thalami (Hedges’ g>0.90). At the global level, PwMS showed significant disruption of the multiplex core-periphery organization (κ=-0.19, Hedges’ g=0.61, *p*<0.001), correlating with clinical phenotype (F=5.42, *p*=0.001), EDSS (rho=-0.08, *p*=0.01) and SDMT (r=0.19, *p*<0.0001). Multiplex κ was the only connectomic measure adding to conventional MRI for the prediction of disease status and cognitive impairment, while physical disability depended also on single-layer contributions.

We show that multilayer networks represent a biologically and clinically meaningful framework to model multimodal MRI data, with disruption of the core-periphery structure emerging as a potential novel biomarker for disease severity and cognitive impairment in multiple sclerosis.

## Introduction

In multiple sclerosis, there is a well-recognized gap between clinical-cognitive impairment and brain pathology as assessed through conventional MRI.^1^ The field of connectomics has now started to bridge this gap, as clinically relevant disruptions to macro-scale brain networks, measured using structural (sMRI), diffusion (dMRI), or resting-state functional (rs-fMRI) MRI, have been extensively demonstrated in people with multiple sclerosis (PwMS), to the point that it has been described as a network disorder.^2, 3^

Our current understanding points towards abnormal connectivity centred around hubs like the thalamus and the default mode network, evolving along the disease course and representing a possible common mechanism through which cumulative brain damage eventually leads to long-term disability.^2, 3^ Nevertheless, MRI-based connectivity studies yield conflicting results, somehow failing to identify a unified connectomic hallmark of multiple sclerosis and related disability.^4, 5^ While this is partly explained by multiple sclerosis’ intrinsic neurobiological and phenotypic heterogeneity,^6, 7^ methodological issues may as well play a role, including the disparity of image processing strategies, and the small sample sizes. Also, one major conceptual problem lies in the focus on single-modality networks, providing only a partial representation of the brain’s complex organization.

Indeed, despite the increasing availability of multimodal neuroimaging data, most studies so far have focused on one aspect of brain connectivity using a single imaging modality (e.g., morphological covariance, MC, with sMRI; structural connectivity, SC, with dMRI; functional connectivity, FC, with rs-fMRI).^2^ Integrating different neuroimaging modalities into a unified brain network model holds promise to enhance our understanding of the brain and its disorders, by informing us about how structure shapes function, how they are jointly impacted by disease, and which aspects are relevant for cognitive functioning and clinical manifestations.^8^ In particular, the brain can be modelled as a multilayer network where different connectivity domains, each encoding a specific type of information about the system, are jointly embodied in the same topological space.^9, 10^ Intriguingly, such a model can capture higher-order emergent properties that are not evident from conventional single-layer architectures, thus potentially containing additional and novel information relating to clinical status.^9–11^

Among the topological properties shown to meaningfully scale to a multilayer setting is the core-periphery structure,^12, 13^ a fundamental feature of many real-world complex networks (including the brain), characterized by a subgraph of densely connected and topologically central nodes (the *core*), and a set of nodes that are strongly connected with the core but sparsely interconnected with each other (the *periphery*).^14^ A strong core-periphery organization is thought to be crucial for healthy brain functioning as it optimizes efficiency-cost balance and robustness to perturbations (all critical constraints on brain network topology),^14^ and its disruption within single connectivity layers has been shown in several psychiatric^15^ and neurological^16^ disorders, including multiple sclerosis.^17^ However, while it has been shown that the core of the human connectome can be more accurately described in a multilayer setting,^12^ the impact of brain pathology on the multilayer core-periphery structure is still largely uncharted.

Here, leveraging unique access to a large multicentric cohort of PwMS, we used a multilayer network approach to integrate information from sMRI, dMRI, and rs-fMRI data and portray an enriched representation of the brain’s core-periphery organization. We hypothesised that joint brain network changes across morphological, structural, and functional levels would reveal a more consistently disrupted multilayer core-periphery structure in PwMS compared to healthy people. A multilayer analysis should be more sensitive to multiple sclerosis-related pathophysiological alterations and enable more accurate predictions of physical and cognitive disability.

## Materials and methods

### Participants

In this retrospective, cross-sectional study, we collected MRI and clinical data of people diagnosed with multiple sclerosis according to 2010 McDonald criteria^18^ or clinically isolated syndrome (CIS)^19^ from 13 European centers (MAGNIMS: www.magnims.eu). Healthy controls (HC) without history of neurologic or psychiatric disorders were also included.

Written informed consent had been obtained from each participant independently at each center. The final protocol for this study was reviewed and approved by the local Ethics Committee and the European MAGNIMS collaboration for the analysis of pseudonymized data.

### Neurological and neuropsychological assessment

At the time of MRI, PwMS were clinically evaluated using the Expanded Disability Status Scale (EDSS)^20^ and the Symbol Digit Modalities Test (SDMT),^21^ measuring physical disability and cognition, respectively. Raw SDMT scores were transformed to age-, sex- and education-adjusted z-scores according to population-specific normative data.^22–26^

### MRI data acquisition and processing

All participants were imaged on 3T scanners with a brain MRI protocol including isotropic T1- weighted (T1w), T2-weighted fluid-attenuated inversion recovery (FLAIR), dMRI, and RS-fMRI sequences. Details of the different acquisition protocols are provided in Supplementary Table 1, while a schematic illustration of the analysis pipelines discussed below is shown in Figure 1.

**Figure 1.**
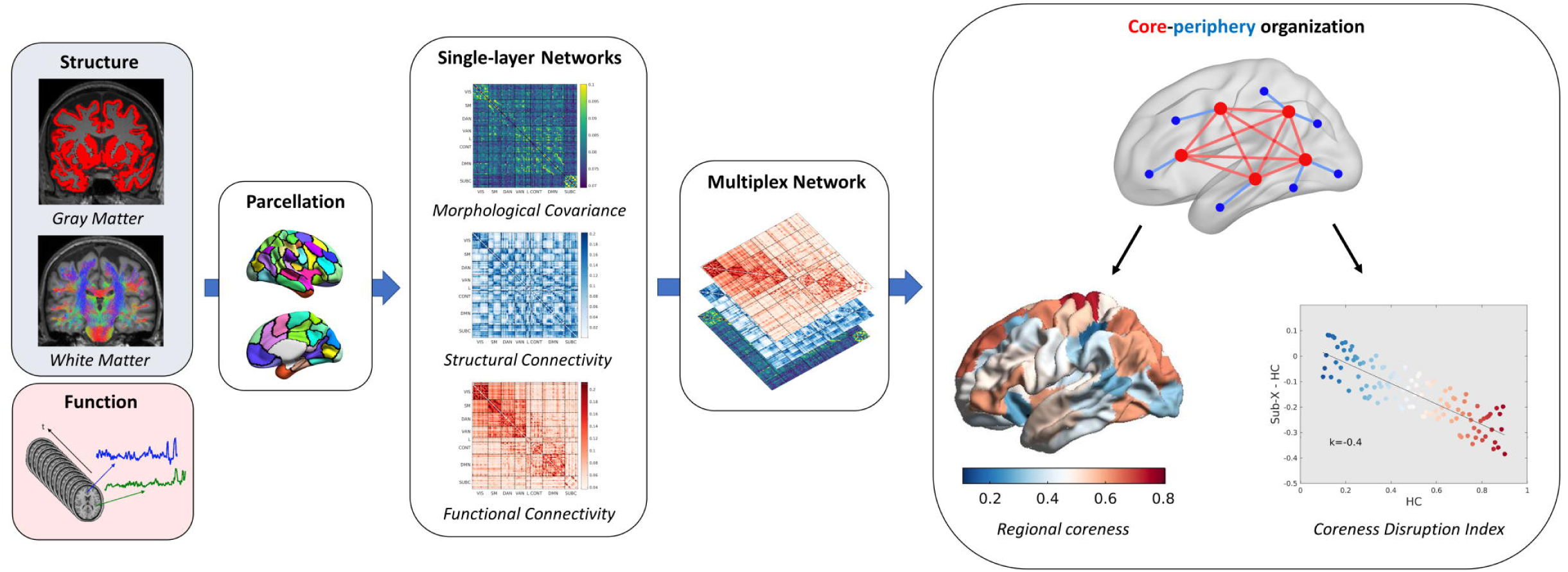
Schematic illustration of the analysis pipeline. SMRI, dMRI and rs-fMRI are processed using the same brain parcellation scheme to obtain networks of morphological covariance, structural connectivity, and functional connectivity, respectively. Connectivity matrices are then merged in a multiplex network, a particular case of multilayer network where there is a one-to-one correspondence between nodes at different layers. The multiplex core- periphery organization is characterized in terms of regional coreness, defined as the probability of a node being part of the multiplex core, and coreness disruption index (κ), quantifying the global weakening of the core-periphery structure.

#### Structural MRI and morphological covariance networks

For PwMS, T2-hyperintense lesions were automatically segmented on FLAIR images using the Lesion Segmentation Tool (LST) 3.0.0 (www.statistical-modelling.de/lst.html). Corresponding masks were used to fill lesions in T1w images with estimated white matter (WM) tissue for subsequent analyses^27^ and to compute total lesion volume (TLV). We used the Computational Anatomy Toolbox (CAT12.7, http://www.neuro.uni-jena.de/cat) to segment T1w volumes into grey matter (GM), WM, and cerebrospinal fluid (CSF), and to parcellate the brain into 100 cortical regions (nodes) from the Schaefer atlas.^28^ This functional parcellation is designed to optimize both local gradient and global similarity measures of the fMRI signal.^28^ The nodes are also associated with 7 canonical functional system labels including visual, somatomotor, dorsal attention, ventral attention, limbic, control, and default mode networks.^29^ We chose the 100 parcel version to best fit the spatial resolution of the available data (Supplementary Table 1) and to reduce the impact of an inaccurate alignment between the atlas and individual scans.^30^ In addition, we used FSL-FIRST to segment 14 subcortical GM regions.^31^ Throughout the diffusion and functional workflows, T1w images were used as reference and underwent additional processing steps, including cortical surface reconstruction with recon-all (FreeSurfer 6.0.1).^32^

Single-subject GM networks were obtained by adapting a previously described pipeline.^33^ Briefly, regional GM volumes were transformed into z-scores while adjusting for the physiological (i.e., estimated in the HC group) effects of age, sex and total intracranial volume (TIV), and a measure of shared deviation from the reference norm was computed for each nodes’ pair to fill a 114 x 114 MC matrix.

#### Diffusion MRI and structural connectivity networks

Preprocessing of diffusion MRI data was performed using QSIPrep 0.14.3,^34^ which is based on Nipype 1.6.1.^35^ MP-PCA denoising as implemented in MRtrix3’s dwidenoise was applied with a 5-voxel window, followed by B1 field inhomogeneity correction using dwibiascorrect from MRtrix3 with the N4 algorithm.^36, 37^ FSL (version 6.0.3)’s eddy was used to correct for head motion and eddy currents.^38^ A deformation field to correct for susceptibility distortions was estimated using available sequences (phase-encoding polarity method,^39^ phase-difference B0 estimation,^40^ or registration-based fieldmap-less estimation^41^) and used to calculate an unwarped b=0 reference for a more accurate co-registration with the anatomical reference. The diffusion-weighted time-series was then resampled to the T1w volume, producing a preprocessed diffusion-weighted series with 2mm isotropic voxels. Then, multi-tissue fiber response functions were generated using the Dhollander algorithm,^42^ and fiber orientation distributions (FODs) were estimated via constrained spherical deconvolution and intensity-normalized using mtnormalize.^43, 44^ Tractography was performed based on WM FODs with MRtrix3’s tckgen, using the iFOD2 probabilistic tracking method to generate 10 million streamlines, with anatomical constraints provided by a hybrid surface/volume segmentation created ad hoc.^45, 46^

Finally, weights for each streamline were calculated using SIFT2^47^ and a 114 x 114 SC matrix was filled with the sums of weights of streamlines connecting each node’s pair. In addition, structural connectivity matrices were log10-transformed to better account for differences at different magnitudes and to make the distribution of edges’ weight more comparable to other layers.^14, 48^

#### Resting-state functional MRI and functional connectivity networks

Preprocessing of rs-fMRI data was performed using fMRIPrep 20.2.6,^49^ which is based on Nipype 1.7.0.^35^ From blood oxygenation level dependent (BOLD) data, a reference volume and its skull-stripped version were generated using a custom methodology of fMRIPrep. Similar to dMRI processing, a deformation field to correct for susceptibility distortions was estimated based on available sequences and used to calculate a corrected EPI reference for a more accurate co-registration with the anatomical reference. The BOLD reference was then co-registered to the T1w reference using bbregister (FreeSurfer) which implements boundary-based registration.^50^ Head-motion parameters for the BOLD reference were estimated before any spatiotemporal filtering using mcflirt (FSL 5.0.9).^51^ After slice-timing correction using 3dTshift from AFNI 20160207,^52^ the BOLD time-series were resampled onto their original, native space by applying a single, composite transform to correct for head-motion and susceptibility distortions. Several confounding time-series were calculated based on the preprocessed BOLD, including the identification of noise components using ICA-AROMA.^53^ Nuisance variables were removed from preprocessed BOLD using Nilearn 0.8.1,^54^ following a previously described strategy^55^ including removal of the first 4 timepoints, band-pass filter (0.008-0.08 Hz), detrending, standardization and confound regression (non-aggressive ICA-AROMA denoising plus removal of mean WM and CSF signal).^53, 56^

Finally, residual mean BOLD time series were obtained from the atlas-defined parcels, and, for each node’s pair, the Pearson correlation coefficient was computed and Fisher z-transformed to fill a 114 x 114 FC matrix. In addition, matrices were absolutized as inverse correlations may encode relevant information and most analysis strategies tend to neglect negative values.^55, 57^

#### Quality control and cross-site harmonization

MRI quality was assessed through metric-guided visual inspection. Scans that were marked as outliers (i.e., outside 1.5 times the interquartile range in the adverse direction of the measurement distribution) according to one or more image quality metrics obtained via CAT12 (for sMRI), qsiprep (for dMRI) and mriqc 0.16.1 (for sMRI and rs-fMRI)^58^ were reviewed and discarded based on visual evaluation where appropriate.

To eliminate nonbiological site-related variability, we used ComBat harmonization to model and remove site effects from brain volumes and structural and functional connectivity matrices, while preserving the biological associations with age, sex and disease status.^59^

### Multiplex networks and core-periphery organization

To correct for differences in average link weight across layers, MC, SC and FC connectivity matrices underwent singular-value decomposition normalization before the construction of a multimodal multiplex, a particular case of multilayer network where there is a one-to-one correspondence between nodes at different layers.^60^ For each brain region, we extracted coreness using the brain network toolbox (https://github.com/brain-network/bnt), following a previously described procedure.^12^ Briefly, each layer is filtered by preserving the strongest weights for the full range of density-based thresholds. At each threshold, a measure of node richness in the multiplex setting is computed by linearly combining node strengths in all layers through a vector of coefficients modulating the relative importance of each layer. We derived the coefficients in a data-driven manner, exploring the parameter space to maximize the separation between PwMS and HC regional coreness. Specifically, we used a Bayesian optimization algorithm implemented in MATLAB R2020a (The MathWorks Inc.), with default parameters, to maximize the ratio of the between-class variance to the within-class variance (i.e., Fisher’s optimization criterion).^13^

Multiplex richness is then fed into a core-periphery decomposition procedure, and coreness is calculated as the number of times that each node is present in the network core across all explored thresholds, normalized by the maximum theoretical value. Coreness disruption index (κ), representing a global measure of core-periphery reorganization, was also computed as the slope of the linear regression model between the mean local coreness of the HC group at each node, taken as a reference, and the differential nodal coreness between that reference and the subject(s) under study.^61^

### Statistical analysis

Second-level analyses were carried out using R (version 4.1.2), and MATLAB (R2020a). The effects of biological confounders (i.e., age and sex) on variables of interest (i.e., regional coreness and coreness disruption index) were removed using nuisance regression, with weights estimated in the HC group. Significance level was set at α=0.05 for all tests, adjusting for multiple comparisons when appropriate.

Differences between patients and HC in terms of κ and nodal coreness (over the full set of brain parcels) were assessed using permutation t tests, controlling for the false discovery rate with the Benjamini-Hochberg procedure.^62^ Additionally, to assess possible κ differences across clinical phenotypes (CIS vs RRMS vs SPMS vs PPMS), we performed a one-way ANOVA analysis, with post hoc tests using Tukey’s method. The associations between κ and clinical measures of physical (EDSS) and cognitive (SDMT) disability were assessed using Spearman and Pearson correlation, respectively.

To evaluate the added value of our multiplex approach over single-layer metrics, we computed κ within the MC, SC, and FC layers separately and looked at the ability of measures in different domains to discriminate between PwMS and HC, as well as between different levels of physical disability (EDSS≥4 vs EDSS<4),^63^ and between impaired (SDMT z-score < -1.5) and preserved information processing speed (IPS-I vs IPS-P).^22^ Comparison was made also with other established MRI measures of multiple sclerosis-related brain damage (i.e., age- and sex-adjusted BPF, and TLV). First, we compared effect sizes (Hedges’ g) of the between-groups differences for the different MRI-derived variables, by computing 95% bootstrap confidence intervals with 5000 resamples. Additionally, we used κ in the different domains and conventional MRI measures to train and validate random forest models for the prediction of disease status (PwMS vs HC), level of physical disability and IPS impairment. Specifically, decision tree learners were combined with bootstrap aggregation, and relevant hyperparameters were tuned using Bayesian optimization in order to minimize the 10-fold CV classification error (1 - accuracy). Model performance was expressed with out-of-bag (OOB) accuracy, while the relative weight of different predictors was estimated using OOB permutation feature importance.^64^

### Data availability

Data from patients are controlled by the respective centers (listed in Supplementary Table 1) and therefore are not publicly available. Request to access raw data should be forwarded to data controllers via the corresponding author. Derived data supporting the findings of this study can be requested by qualified investigators from the corresponding author.

## Results

### Participants

A total of 1517 participants were considered for this study. Of these, 33 were excluded due to poor MRI quality or image processing failure, leading to a final population including 1048 PwMS and 436 HC. Demographic, clinical, and conventional MRI characteristics are reported in Table 1.

**Table 1.**
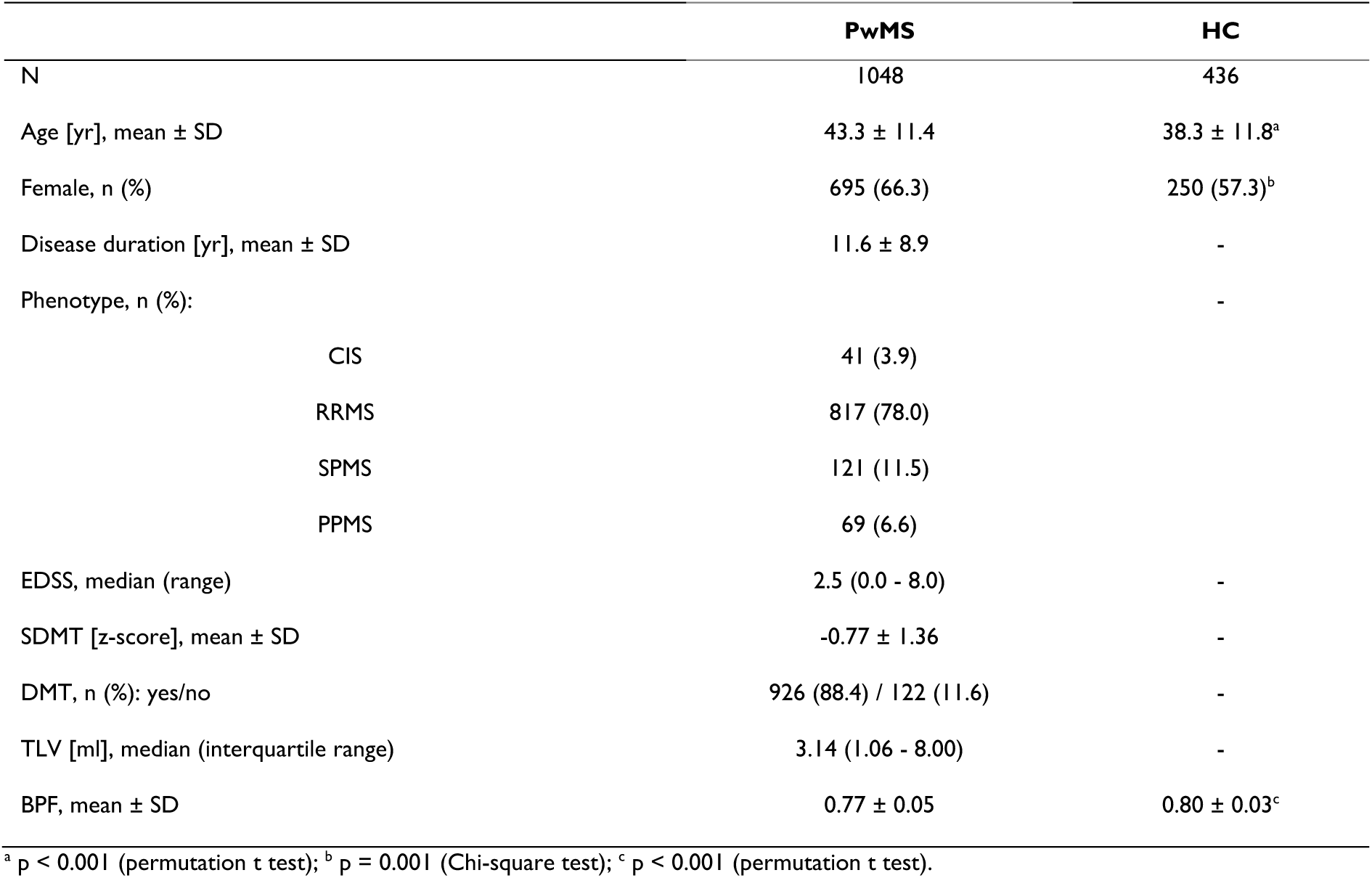
Demographic, clinical, and MRI characteristics of the studied population. SD = standard deviation; PwMS = people with multiple sclerosis; CIS = clinically isolated syndrome; RRMS = relapsing-remitting multiple sclerosis; SPMS = secondary-progressive multiple sclerosis; PPMS = primary-progressive multiple sclerosis; HC = healthy controls; EDSS = Expanded Disability Status Scale; SDMT = Symbol Digit Modalities Test; DMT = disease-modifying treatment; TLV = total lesion volume; BPF = brain parenchymal fraction.

### Multiplex core-periphery organization

Average preprocessed connectivity matrices in the HC group are shown in Supplementary Figure 1. Following the optimization procedure, the coefficients maximizing the separation between PwMS and HC in terms of regional coreness were determined to be 0.75, 0.79, and 0.24 for the MC, SC, and FC layers, respectively.

In the HC group, the multiplex core included on average subcortical GM structures, especially the thalami and putamina, as well as cortical areas participating in both sensorimotor (somatomotor and visual) and associative (default-mode, control, and attention) networks (Figure 2). Maps of average HC regional coreness in the single-layer domains are shown in Supplementary figure 2.

**Figure 2.**
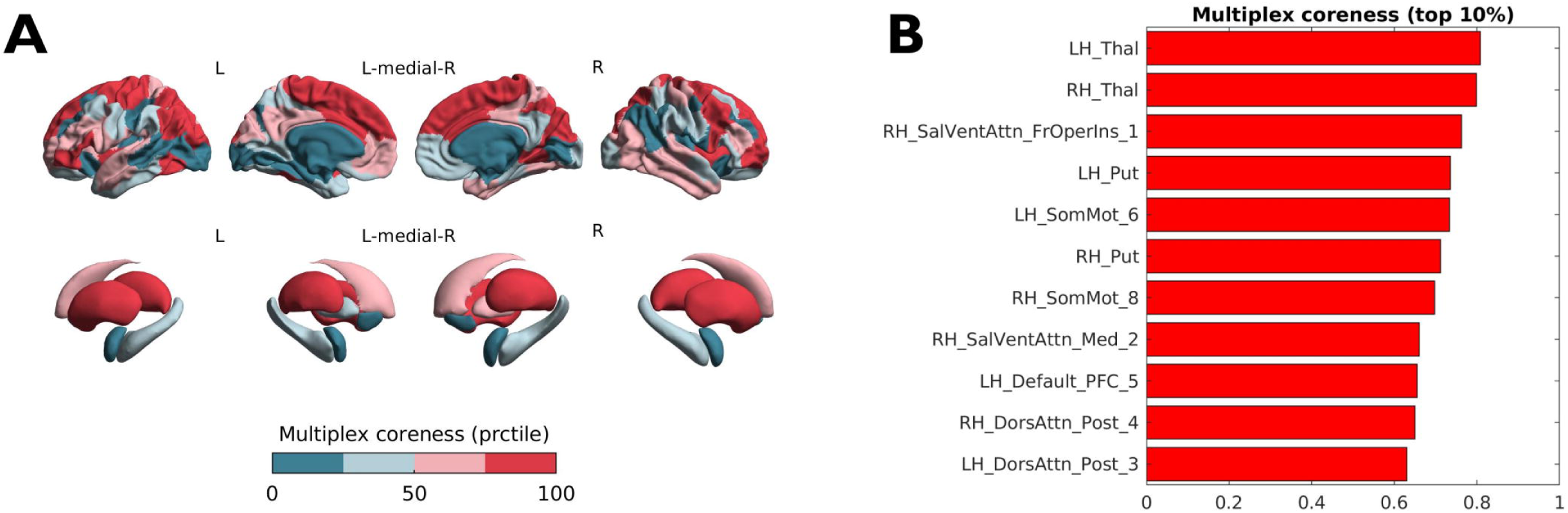
Average multiplex coreness in the healthy controls group. (**A**) Color-coded (*teal* to *red*) map of multiplex coreness percentile ranks superimposed on surface renderings of the cortex and subcortical structures. Image was obtained with the ENIGMA toolbox.^80^ (**B**) Highest 10% multiplex coreness nodes and corresponding absolute values are shown. Nomenclature of cortical areas follows the 7-network Schaefer-100 parcellation.^28^

### Disrupted multiplex core-periphery structure in multiple sclerosis

PwMS showed widespread deviations in regional coreness compared to the HC group (Figure 3A), with the greatest effect sizes observed at the level of deep GM structures (reduced coreness) and associative areas in the medial prefrontal, cingulate and lateral temporal cortices (increased coreness) (Supplementary Table 2). At the global level, the anatomical distribution of the observed changes was such that topologically central nodes were generally more impacted than peripheral ones (which tended to have preserved or even increased coreness values), as expressed by κ = -0.19 (Hedges’ g = 0.61 *p* < 0.001) (Figure 3B).

**Figure 3.**
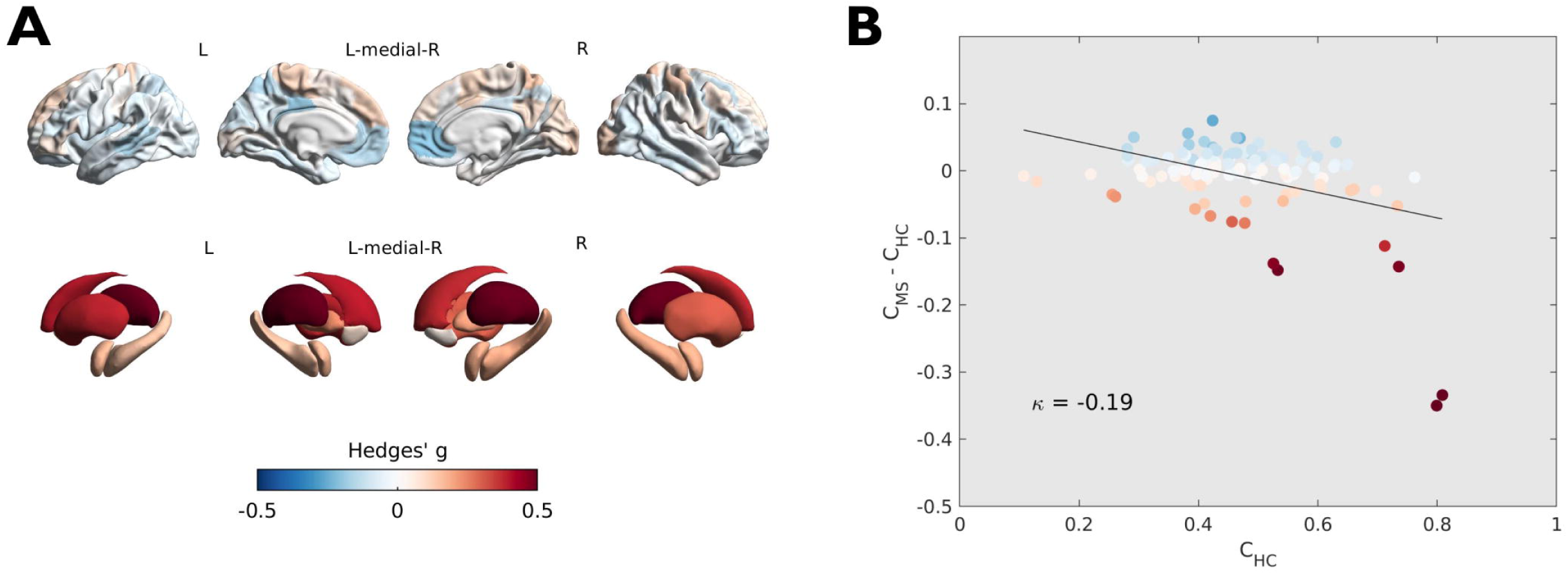
Differences in regional coreness between PwMS and HC and coreness disruption index. (**A**) Color-coded (*blue* to *red*) map of effect sizes (Hedges’ g) of the between-group difference superimposed on surface renderings of the cortex and subcortical structures. Image was obtained with the ENIGMA toolbox.^80^ (**B**) Scatterplot showing the between-group difference in regional coreness as a function of the average coreness in the HC group. The slope of the linear regression line corresponds to the coreness disruption index κ = -0.19. Each circle represents a brain region, color-coded as in panel A.

There was a significant effect of clinical phenotype on the weakening of the core-periphery structure of multimodal brain networks (F[3, 1044] = 5.42, *p* = 0.001), with progressively greater disruption going from CIS to SPMS participants, and intermediate κ values in patients with PPMS (Figure 4). Moreover, patients with a stronger core-periphery organization had lower EDSS (rho = -0.08, *p* = 0.01) and higher SDMT (r = 0.19, *p* < 0.0001) scores (Figure 5).

**Figure 4.**
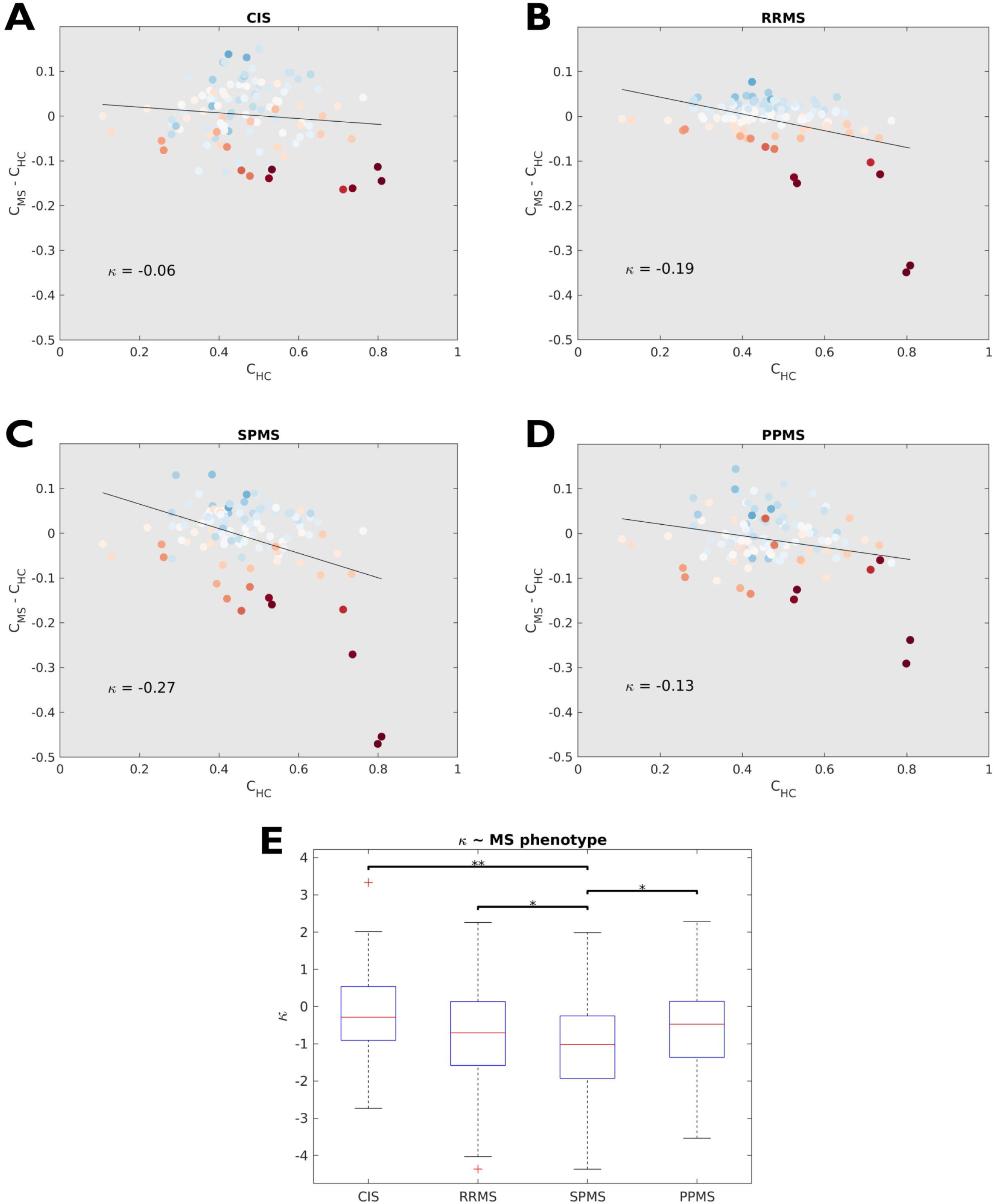
Coreness disruption index and clinical phenotypes. Coreness disruption index (κ) plots are shown for (**A**) clinically isolated syndrome (CIS), (**B**) relapsing-remitting (RRMS), (**C**) secondary-progressive (SPMS), and (**D**) primary-progressive (PPMS) patients. (**E**) Boxplots showing the distributions of κ values across different phenotypes. (*) Adjusted *p* < 0.05; (**) Adjusted *p* < 0.01. In (**A-D**), each circle represents a brain region, with color encoding the magnitude of the between-group (MS/CIS versus HC) difference in terms of regional coreness.

**Figure 5.**
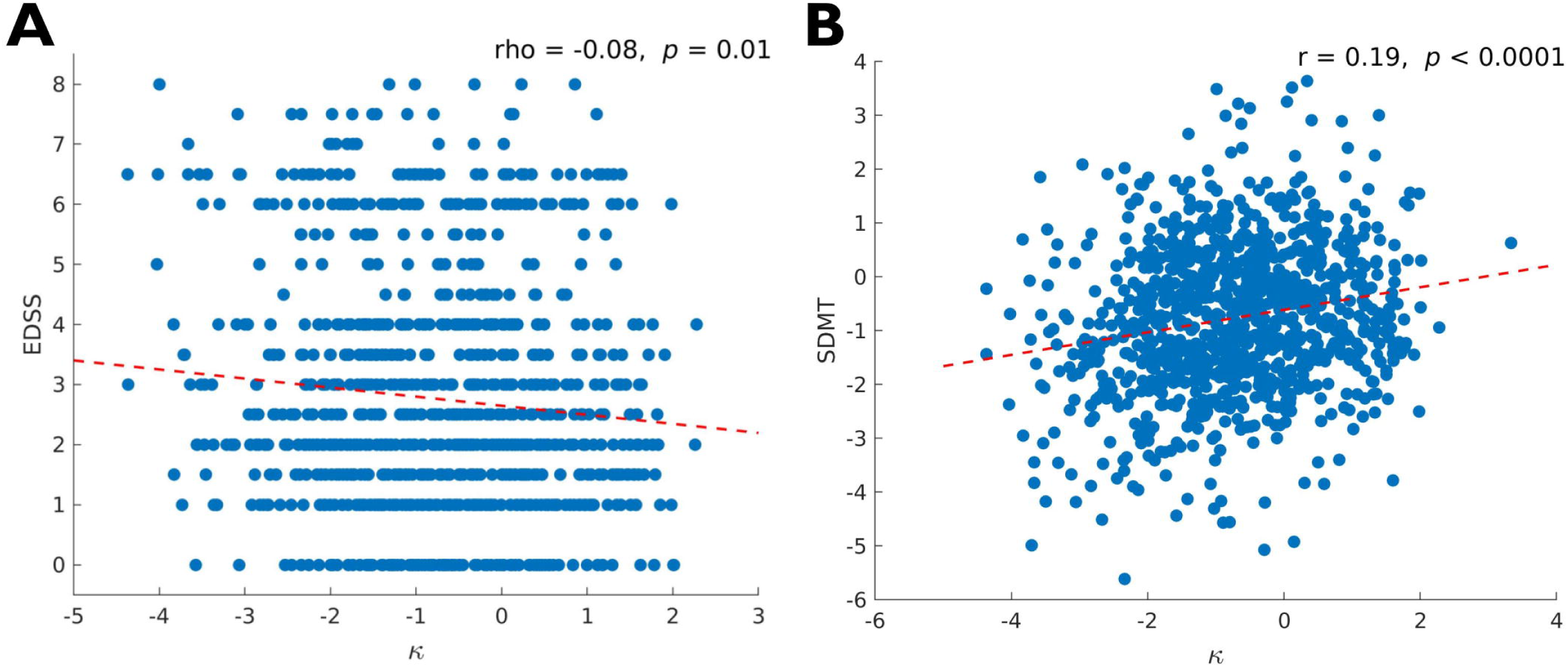
Coreness disruption index and clinical variables. Scatterplots showing the correlations of coreness disruption index (κ) with (**A**) EDSS and (**B**) SDMT scores. Both κ and SDMT are expressed as confounder-adjusted z-scores. EDSS = Expanded Disability Status Scale; SDMT = Symbol Digit Modalities Test.

### Added value of multiplex over single-layer network measures

Of all PwMS, 27 % had an EDSS score ≥ 4 and 29 % had impaired IPS on the SDMT. Disease status (CIS/MS vs HC) and IPS performance (IPS-I vs IPS-P) were more strongly associated with disruption of the core-periphery structure in the multiplex setting (Hedges’g = 0.61 [95% CI = 0.51 – 0.72] and 0.40 [95% CI = 0.26 – 0.54], respectively) than with any of the single-layer measures. As for the level of physical disability (EDSS≥4 vs EDSS<4), the effect size associated with the disruption of the multiplex core-periphery organization (Hedges’ g = 0.13 [95% CI = 0.00 – 0.25]) was not significantly higher than for homologous single-layer measures (Figure 6A-C).

**Figure 6.**
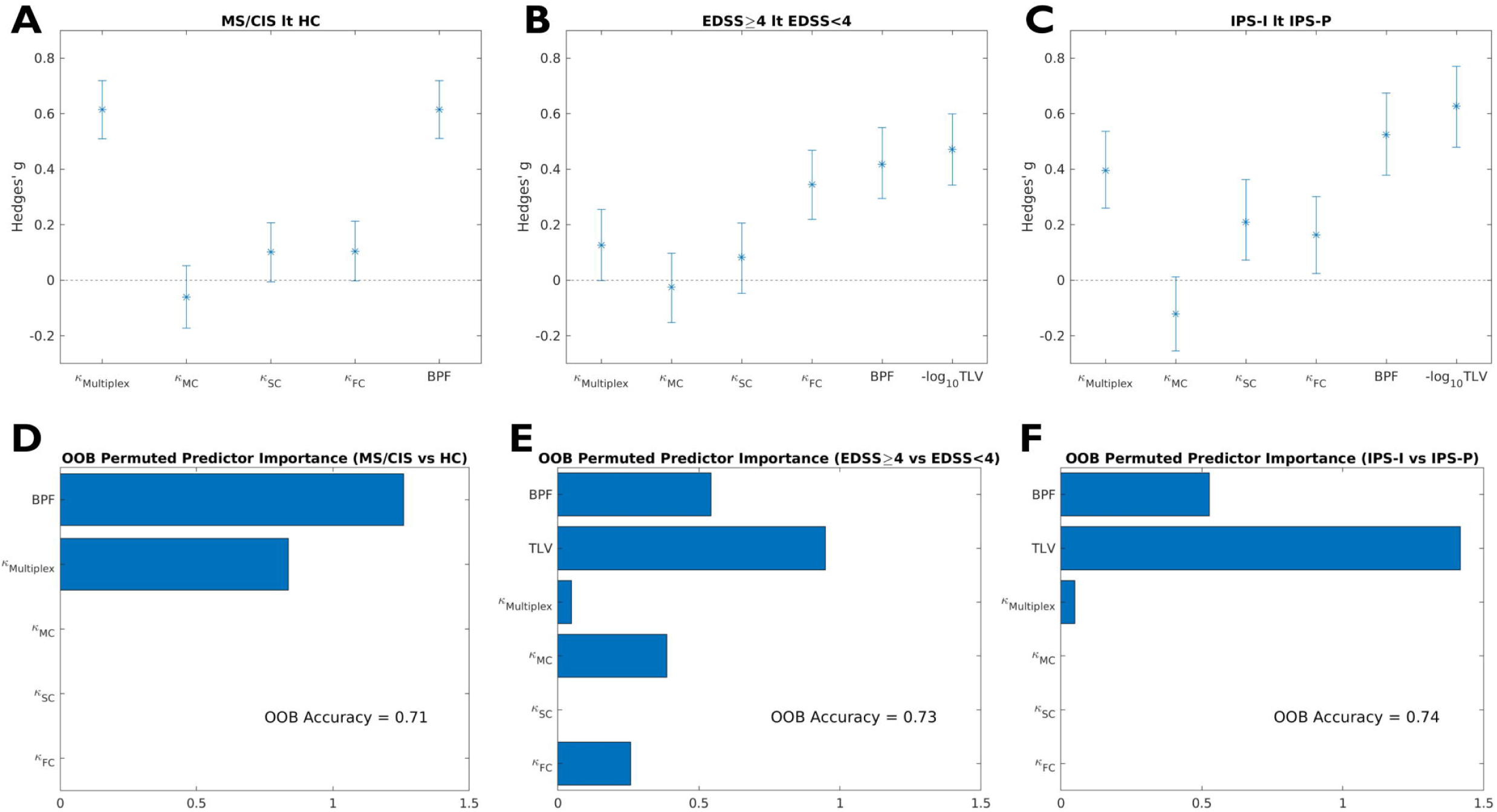
Added value of multiplex over single-layer measures. For indeces of coreness disruption in the multiplex and single-layer domains, as well as for brain parenchymal fraction (BPF) and total lesion volume (TLV), shown are: (*top row*) the effect sizes (Hedges’ g) and corresponding 95% confidence intervals associated with differences between (**A**) PwMS and healthy controls (MS/CIS vs HC), (**B**) patients with high and low levels of physical disability (EDSS≥4 vs EDSS<4), and (**C**) patients with impaired and preserved information processing speed (IPS-I vs IPS-P); (*bottom row*) results of the random forest classifiers and corresponding predictor importance analyses for the prediction of (**D**) disease status (MS/CIS vs HC), (**E**) level of physical disability (EDSS≥4 vs EDSS<4), and (**F**) impaired information processing speed (IPS-I vs IPS-P).

Random forest models leveraging both conventional and global core-periphery organization-related MRI measures reached OOB accuracies of 0.71, 0.73 and 0.74 for the CIS/MS vs HC, EDSS≥4 vs EDSS<4 and IPS-I vs IPS-P classifications, respectively. For both disease and cognitive status predictive models, disruption of the core-periphery structure in the multiplex domain was the only connectomic metric independently contributing to the classification along with MRI-derived volumes. On the other hand, core-periphery disruption in single-layer domains (i.e., MC and FC) were at least as important as the homologous multiplex measure in predicting the level of physical disability (Figure 6D-F).

## Discussion

By jointly modelling three different MRI modalities in a multilayer framework, we revealed clinically relevant disruption of the core-periphery organization of multimodal (structural-functional) brain networks in a large multicentric sample of PwMS. We showed that the degree of weakening of the multiplex core-periphery depends on the disease phase and is associated with physical disability and cognition, being more sensitive than conventional single-layer measures to multiple sclerosis-related pathophysiological and cognitive changes.

As the information conveyed by connectivity data is multivariate in nature and multimodal datasets become increasingly available, it has been advocated that multilayer networks, rather than single-layer architectures, may represent the ideal mathematical framework to study the brain as a complex system.^10–12^ However, how to meaningfully model together structural and functional aspects of brain connectivity is still debated, with novel possible methodological solutions continuing to emerge.^10–12^

The method adopted here to detect the core-periphery of multiplex networks has the advantage of minimizing the need for a priori assumptions, reducing the variable degree of arbitrariness and information loss that are inevitably associated with the processes of, e.g., thresholding/binarizing connectivity matrices or assigning predefined weights to the different layers.^12^ In our large multi-centre population, the data-driven optimization procedure showed that all three modalities are necessary to maximize the separation between PwMS and HC in regional coreness, with greater estimated contribution coefficients for structural rather than functional layers. This is in line with previous findings in Alzheimer’s disease,^13^ and may be partially explained by the known robustness of diffusion-based networks relative to functional layers, which are characterized by higher inter-subject variability.^65, 66^ Also, this is consistent with the interpretation of multiple sclerosis being primarily characterised by cumulative structural brain damage, with functional organization that can be relatively preserved or exhibit compensatory changes, especially in the early phases.^67^

In keeping with previous knowledge on the structural and functional cores of the human connectome,^68–70^ average coreness maps in the HC group revealed that the SC core included the superior frontal and superior parietal cortex, as well as subcortical GM structures, while rolandic and occipital cortical regions participating in the somatomotor and visual networks constituted the functional core. On the other hand, a less pronounced core-periphery organization was observed in the MC layer, with lower absolute values of core nodes resulting from a more distributed coreness pattern. Notably, while the coreness of the multiplex network was strongly influenced by the SC layer, it also captured the role of functional hubs (e.g., in the occipital cortices) whose importance was neglected by diffusion-based networks. This confirms again that, while being sensitive to single-layer contributions, the multiplex setting provides a unique representation of the brain core-periphery structure.^12^

In PwMS, the regional coreness profile deviated extensively from the control group, with some increases in the associative cortex, a prominent decrease in subcortical GM structures, and the greatest effect sizes observed at the level of the thalami. Indeed, the thalamus is widely recognized as a vulnerable site for multiple sclerosis-related damage, with atrophy, structural disconnection and functional reorganization occurring from the early stages, evolving with the disease course and driving disability progression and cognitive impairment.^71^ Hence, it is not surprising to observe how thalamic structural and functional modifications result in a reduced topological centrality in the multiplex setting.

At the global level, multiple sclerosis was associated with weakening of the multiplex core-periphery structure, with hub regions found to be more impacted than would be expected based solely on their reference coreness in non diseased subjects. From a network science perspective, this conceptually equates multiple sclerosis with a targeted (i.e., network elements are impacted according to some index of topological centrality), rather than a random (i.e., network elements are impacted with uniform probability), attack.^72^ Previous evidence from sMRI,^73^ dMRI,^17^ and fMRI^2^ studies suggested that multiple sclerosis-related brain damage occurs in a non-random, network-mediated fashion, a hypothesis that bears great transdiagnostic relevance as it seems to apply to many different neuropsychiatric disorders.^74, 75^ Several mechanisms (not necessarily mutually exclusive) have been proposed to explain this phenomenon, including diaschisis/transneuronal degeneration, nodal stress, shared vulnerability, and propagation of toxic agents/neuroinflammatory response along neuronal connections.^76, 77^ Our multimodal analysis suggests that these processes are likely to impact the different layers of brain connectivity in a synergistic manner, as witnessed by the greater sensitivity of the multimodal network model towards multiple sclerosis-related changes compared to single-layer architectures.

While our sample was largely composed of patients with relapsing-remitting multiple sclerosis, an association between clinical phenotype and weakening of the core-periphery structure was still observable, with a linear trend of progressive disruption in relapse-onset forms suggesting that it may parallel the progression of brain damage along the disease course. Also, disruption of the multiplex core-periphery organization significantly explained physical and cognitive disability, in keeping with the idea of the core-periphery structure as a fundamental organizing principle of brain networks, supporting efficient information integration and ultimately healthy brain functioning.^14^

Core-periphery disruption in multimodal, rather than single-layer, networks contributed to the prediction of cognitive impairment, adding to conventional MRI measures and confirming that the information conveyed by multilayer networks is more than just the sum of its parts, capturing emergent properties that are relevant for cognition but not evident from single layers (alone or in combination). On the other hand, a noisier picture emerged for the prediction of physical disability, although the known limitations of EDSS from the clinimetric point of view and the contribution of spinal cord damage (unexplored here) are likely to play an important _role._78,79

The present study is not without limitations. First, the proposed approach is only one of the many possible solutions to model multivariate brain connectivity data, with alternative methods that may be more appropriate according to the research question and the available data or resources. Also, the purely cross-sectional nature of our dataset limits the potential for investigating causal relationships and exploring the prognostic value of the observed connectomic changes. In addition, clinical evaluations were limited to only EDSS and SDMT, while more advanced measures (e.g., assessing additional cognitive domains and specific motor functions) could yield additional information. Finally, we used multilayer networks with the merely descriptive purpose of characterizing brain connectivity modifications and their clinical correlates in multiple sclerosis. Future studies following a predictive approach will be needed to drive the analysis of multimodal connectivity data towards translational clinical impact.

In conclusion, we show that multilayer networks represent a biologically and clinically meaningful framework to jointly model multimodal MRI data, with disruption of the core-periphery structure emerging as a potential novel biomarker for disease severity and cognitive impairment in multiple sclerosis.

## Acknowledgements

Jérémy Guillon, PhD, is gratefully acknowledged for the original implementation of the code for the analysis of the core-periphery structure of multiplex networks (https://github.com/brain-network/bnt).

## Funding

G.P. was supported by the ECTRIMS-MAGNIMS Research Fellowship Programme (2020).

F.P. and B.K. are supported by the UK National Institute for Health Research (NIHR) Biomedical Research Centre (BRC) at UCLH and UCL. A.C. is supported by EUROSTAR E!113682 HORIZON2020. Sa.Co. is supported by a Rosetrees Trust Grant (PGL21/10079).

M.A.F. is supported by a grant from the MRC (MR/S026088/1). L.D. Is supported by the Dutch Research Council (NWO, Vidi 198.015). The study was supported by grants from The Research Council of Norway (NFR, grant number 240102) and the South-Eastern Health Authorities of Norway (grant number 257955).

## Competing interests

M.C. received speaker honoraria from Biogen, Bristol Myers Squibb, Celgene, Genzyme, Merck Serono, Novartis, and Roche and receives research support from the Progressive MS Alliance and Italian Minister of Health. Si.Co. serves on scientific advisory board for Amicus Therapeurics, has received speaker honoraria from Sanofi and research grants from Fondazione Italiana Sclerosi Multipla and Telethon. R.S. was awarded a MAGNIMS-ECTRIMS fellowship in 2019. M.F. is Editor-in-Chief of the Journal of Neurology, Associate Editor of Human Brain Mapping, Neurological Sciences, and Radiology; received compensation for consulting services from Alexion, Almirall, Biogen, Merck, Novartis, Roche, Sanofi; speaking activities from Bayer, Biogen, Celgene, Chiesi Italia SpA, Eli Lilly, Genzyme, Janssen, Merck-Serono, Neopharmed Gentili, Novartis, Novo Nordisk, Roche, Sanofi, Takeda, and TEVA; participation in Advisory Boards for Alexion, Biogen, Bristol-Myers Squibb, Merck, Novartis, Roche, Sanofi, Sanofi-Aventis, Sanofi-Genzyme, Takeda; scientific direction of educational events for Biogen, Merck, Roche, Celgene, Bristol-Myers Squibb, Lilly, Novartis, Sanofi-Genzyme; he receives research support from Biogen Idec, Merck-Serono, Novartis, Roche, Italian Ministry of Health, Fondazione Italiana Sclerosi Multipla, and ARiSLA (Fondazione Italiana di Ricerca per la SLA). The University Hospital Basel (USB), as the employer of C.G., has received the following fees which were used exclusively for research support: (i) advisory board and consultancy fees from Actelion, Genzyme-Sanofi, Novartis, GeNeuro and Roche; (ii) speaker fees from Genzyme-Sanofi, Novartis, GeNeuro and Roche; (iii) research support from Siemens, GeNeuro, Roche. C.G. is supported by the Swiss National Science Foundation (SNSF) grant PP00P3_176984, the Stiftung zur Förderung der gastroenterologischen und allgemeinen klinischen Forschung and the EUROSTAR E!113682 HORIZON2020. E.A.H. received honoraria for lecturing and advisory board activity from Biogen, Merck and Sanofi-Genzyme and unrestricted research grant from Merck. S.L. received compensation for consulting services and speaker honoraria from Biogen Idec, Novartis, TEVA, Genzyme, Sanofi and Merck. S.M. received honoraria for lecturing and advisory board activity from UCB and Biogen, and travel grant from Roche and Merck. M.M. has received research grants from the ECTRIMS-MAGNIMS, the UK MS Society, and Merck, and honoraria from Biogen, BMS Celgene, Ipsen, Merck, Novartis and Roche. J.P. has received support for scientific meetings and honorariums for advisory work From Merck Serono, Novartis, Chugai, Alexion, Roche, Medimmune, Argenx, UCB, Mitsubishi, Amplo, Janssen, Sanofi. Grants from Alexion, Roche, Medimmune, UCB, Amplo biotechnology. Patent ref P37347WO and licence agreement Numares multimarker MS diagnostics Shares in AstraZenica. Acknowledges Partial funding by Highly specialised services NHS England. M.P. discloses travel/meeting expenses from Novartis, Janssen, Roche and Merck, speaking honoraria from HEALTH&LIFE S.r.l., honoraria for consulting services from Biogen and research grants from Baroni Foundation. D.P. has received funding for travel from Merck, Genzyme/Sanofi-Aventis and Biogen, as well as speaking honoraria from Biogen, Novartis and Merck. M.A.R. received consulting fees from Biogen, Bristol Myers Squibb, Eli Lilly, Janssen, Roche; and speaker honoraria from Bayer, Biogen, Bristol Myers Squibb, Bromatech, Celgene, Genzyme, Merck Healthcare Germany, Merck Serono SpA, Novartis, Roche, and Teva. She receives research support from the MS Society of Canada and Fondazione Italiana Sclerosi Multipla. She is Associate Editor for Multiple Sclerosis and Related Disorders. A.T. has been supported by grants from MRC (MR/S026088/1), NIHR BRC (541/CAP/OC/818837) and RoseTrees Trust (A1332 and PGL21/10079), has had meeting expenses from Merck, Biomedia and Biogen Idec and was UK PI for two clinical trials sponsored by MEDDAY (MS-ON - NCT02220244 and MS-SPI2 - NCT02220244). P.V. received speaker honoraria from Biogen Idec. O.C. is an NIHR Research Professor (RP-2017-08-ST2-004); acts as a consultant for Biogen, Merck, Novartis, Roche, and Teva; and has received research grant support from the MS Society of Great Britain and Northern Ireland, the NIHR UCLH Biomedical Research Centre, the Rosetree Trust, the National MS Society, and the NIHR-HTA. M.M.S. serves on the editorial board of Neurology and Frontiers in Neurology, receives research support from the Dutch MS Research Foundation, Eurostars-EUREKA, ARSEP, Amsterdam Neuroscience, MAGNIMS and ZonMW and has served as a consultant for or received research support from Atara Biotherapeutics, Biogen, Celgene/Bristol Meyers Squibb, Genzyme, MedDay and Merck. F.B.: Steering committee and iDMC member for Biogen, Merck, Roche, EISAI. Consultant for Roche, Biogen, Merck, IXICO, Jansen, Combinostics. Research agreements with Novartis, Merck, Biogen, GE, Roche. Co-founder and share-holder of Queen Square Analytics LTD. The remaining authors report no competing interests.

## Supplementary material

Supplementary material is available at *Brain* online.

## Appendix

Authors are members of the MAGNIMS network (Magnetic Resonance Imaging in multiple sclerosis; https://www.magnims.eu/), which is a group of European clinicians and scientists with an interest in undertaking collaborative studies using MRI methods in multiple sclerosis, independent of any other organization. The group is run by a steering committee whose members are: Frederik Barkhof (Amsterdam), Nicola de Stefano (Siena), Jaume Sastre-Garriga (Barcelona, Co-Chair), Olga Ciccarelli (London), Christian Enzinger (Graz), Massimo Filippi (Milan), Claudio Gasperini (Rome), Ludwig Kappos (Basel), Jacqueline Palace (Oxford), Hugo Vrenken (Amsterdam), Alex Rovira (Barcelona), Maria Assunta Rocca (Milan, Co-Chair), and Tarek Yousry (London).

## Notes

### Author Declarations

Ethics Committee of University College London gave approval for this work.

