## Supplementary Material for "More than the sum of its parts: disrupted core-periphery of multiplex networks in multiple sclerosis"

### Supplementary Table 1. MRI acquisition protocols.

| **Centre** | Amsterdam | | | Barcelona | | | Basel | | | Graz | | |
|---|---|---|---|---|---|---|---|---|---|---|---|---|
| **Vendor, Model** | GE, Signa | | | Siemens, Trio | | | Siemens, Prima | | | Siemens, Prisma | | |
| **Years of recruitment** | 2008-2012 | | | 2016-2019 | | | 2019-2021 | | | 2021-2022 | | |
| **Modality** | **3D T1w** | **dMRI** | **rs-fMRI** | **3D T1w** | **dMRI** | **rs-fMRI** | **3D T1w** | **dMRI** | **rs-fMRI** | **3D T1w** | **dMRI** | **rs-fMRI** |
| **Voxel dimensions (mm)** | 0.9x0.9x1 | 2x2x2.4 | 3.3x3.3x3 | 0.94x0.94x0.94 | 1.5x1.5x1.51 | 3x3x3 | 1x1x1 | 1.8x1.8x1.8 | 2x2x2 | 1x1x1 | 1.5x1.5x1.5 | 2x2x2 |
| **TR (ms)** | 7.8 | 13000 | 2200 | 1800 | 14800 | 2000 | 5000 | 4500 | 768 | 1900 | 3318 | 1000 |
| **TE (ms)** | 3 | 91 | 35 | 3 | 103 | 19 | 3 | 75 | 37 | 2.7 | 87.4 | 35 |
| **TI (ms)** | 450 | - | - | 900 | - | - | - | - | - | 900 | - | -- |
| **FA (°)** | 12 | 90 | 80 | 9 | 90 | 90 | | 90 | 52 | 9 | 78 | 70 |
| **Slices, Orientation** | , sagittal | 53, axial | , axial | 240, sagittal | 100, axial | 40, axial | 176, sagittal | 80, axial | 72, axial | 176, sagittal | 96, axial | 54, axial |
| **Directions/Time points** | - | 30 | 202 | - | 60 | 450 | - | 137 | | - | 96 | 300 |
| **b-values (s/mm$^2$)** | - | 1000 | - | - | 1000 | - | - | 700, 1000, 2000, 3000 | - | - | 800, 1600, 2500 | - |
| **EPI distortion correction** | - | Fieldmap-less | Fieldmap-less | - | Phase-difference | Fieldmap-less | - | PEPOLAR | PEPOLAR | - | PEPOLAR | Fieldmap-less |
| | **Number of participants** | | | **Number of participants** | | | **Number of participants** | | | **Number of participants** | | |
| **HC** | 95 | | | 8 | | | 15 | | | 50 | | |
| **CIS** | - | | | - | | | - | | | 3 | | |
| **RRMS** | 238 | | | 51 | | | 81 | | | 127 | | |
| **SPMS** | 51 | | | 9 | | | 3 | | | 10 | | |
| **PPMS** | 36 | | | - | | | 3 | | | 3 | | |
| **QC exclusions** | 8 | | | 1 | | | 1 | | | 1 | | |

| Centre | London | | | Mainz | | | Milan | | | Naples I | | |
|---|---|---|---|---|---|---|---|---|---|---|---|---|
| **Vendor, Model** | Philips, Achieva | | | Siemens, Trio | | | Philips, Ingenia | | | Siemens, Trio | | |
| **Years of recruitment** | 2019-2021 | | | 2017-2019 | | | 2017-2020 | | | 2016-2018 | | |
| **Modality** | **3D T1w** | **dMRI** | **rs-fMRI** | **3D T1w** | **dMRI** | **rs-fMRI** | **3D T1w** | **dMRI** | **rs-fMRI** | **3D T1w** | **dMRI** | **rs-fMRI** |
| **Voxel dimensions (mm)** | 1x1x1 | 2x2x2 | 3x3x3 (0.5-mm gap) | 1x1x1 | 2.5x2.5x2.5 | 3x3x2 (1-mm gap) | 1x1x1 | 2.3x2.3x2.3 | 2.5x2.5x3 | 0.8x0.8x0.8 | 2.2x2.2x2.2 | 3x3x4 (1-mm gap) |
| **TR (ms)** | 7 | 6079 | 4000 | 1900 | 9000 | 3060 | 7 | 5900 | 1560 | 3000 | 7400 | 2500 |
| **TE (ms)** | 3.2 | 96 | 25 | 2.5 | 102 | 30 | 3.2 | 78 | 35 | 2.4 | 88 | 50 |
| **TI (ms)** | - | - | - | 900 | - | - | 1000 | - | - | 1000 | - | - |
| **FA (°)** | 8 | 90 | 90 | 9 | 90 | 90 | 8 | 90 | 70 | 9 | 90 | 90 |
| **Slices, Orientation** | 176, sagittal | 72, axial | 43, axial | 192, sagittal | 62, axial | 49, axial | 204, sagittal | 56, axial | 48, axial | 224, sagittal | 60, axial | 30, axial |
| **Directions/Time points** | - | 76 | 100 | - | 30 | 205 | - | 96 | 320 | - | 64 | 200 |
| **b-values (s/mm$^2$)** | - | 1000, 2000, 2800 | - | - | 900 | - | - | 700, 1000, 2850 | - | - | 1000 | - |
| **EPI distortion correction** | - | PEPOLAR | Fieldmap-less | - | Fieldmap-less | Fieldmap-less | - | PEPOLAR | Fieldmap-less | - | Fieldmap-less | Fieldmap-less |
| | **Number of participants** | | | **Number of participants** | | | **Number of participants** | | | **Number of participants** | | |
| **HC** | 16 | | | 55 | | | 35 | | | 52 | | |
| **CIS** | 34 | | | - | | | - | | | - | | |
| **RRMS** | 25 | | | 50 | | | 29 | | | 30 | | |
| **SPMS** | - | | | - | | | 26 | | | 15 | | |
| **PPMS** | - | | | - | | | 7 | | | 7 | | |
| **QC exclusions** | - | | | 3 | | | 3 | | | 1 | | |

| Centre | Naples II | | | Oslo | | | Oxford | | | Siena | | | Verona | | |
|---|---|---|---|---|---|---|---|---|---|---|---|---|---|---|---|
| **Vendor, Model** | GE, Discovery | | | GE, Discovery | | | Siemens | | | Philips, Achieva | | | Philips, Achieva | | |
| **Years of recruitment** | 2019-2022 | | | 2016-2019 | | | 2018-2019 | | | 2017-2021 | | | 2015-2017 | | |
| **Modality** | **3D T1w** | **dMRI** | **rs-fMRI** | **3D T1w** | **dMRI** | **rs-fMRI** | **3D T1w** | **dMRI** | **rs-fMRI** | **3D T1w** | **dMRI** | **rs-fMRI** | **3D T1w** | **dMRI** | **rs-fMRI** |
| **Voxel dimensions (mm)** | 1x1x1 | 2x2x2 | 2.3x2.3x3 | 1x1x1 | 2x2x2 | 3x3x3 (1-mm gap) | 1x1x1 | 2x2x2 | 2.4x2.4x2.4 | 1x1x1 | 2.5x2.5x2.5 | 1.9x1.9x4 | 1x1x1 | 2x2x2 | 1.8x1.8x4 |
| **TR (ms)** | 7.0 | 7302 | 1500 | 8.2 | 8150 | 2250 | 2040 | 3600 | 735 | 10 | 7053.1 | 3000 | 8.2 | 9300 | 2600 |
| **TE (ms)** | 3.0 | 79.2 | 19 | 3.2 | 83.1 | 30 | 4.7 | 92 | 39 | 4 | 96.4 | 35 | 3.8 | 109 | 35 |
| **TI (ms)** | 650 | - | - | 450 | - | - | 900 | - | - | - | - | - | - | - | - |
| **FA (°)** | 9 | 90 | 90 | 12 | 90 | 79 | 8 | 78 | 52 | 8 | 90 | 90 | 8 | 90 | 90 |
| **Slices, Orientation** | 206, sagittal | 66, axial | 44, axial | 188, sagittal | 67, axial | 43, axial | 192, sagittal | 72, axial | 64, axial | 256, sagittal | 50, axial | 30, axial | 180, sagittal | 62, axial | 35, axial |
| **Directions/Time points** | - | 64 | 320 | - | 60 | 200 | - | 100 | 490 | - | 32 | 200 | - | 96 | 225 |
| **b-values (s/mm$^2$)** | - | 2000 | - | - | 1000 | - | - | 1000, 2000 | - | - | 900 | - | - | 700, 2000 | - |
| **EPI distortion correction** | - | PEPOLAR | Fieldmap-less | - | PEPOLAR | Phase-difference | - | PEPOLAR | Fieldmap-less | - | Fieldmap-less | Fieldmap-less | - | PEPOLAR | Fieldmap-less |
| | **Number of participants** | | | **Number of participants** | | | **Number of participants** | | | **Number of participants** | | | **Number of participants** | | |
| **HC** | 18 | | | 24 | | | 17 | | | 30 | | | 21 | | |
| **CIS** | 1 | | | - | | | - | | | - | | | 3 | | |
| **RRMS** | 51 | | | 56 | | | 16 | | | 92 | | | 44 | | |
| **SPMS** | 5 | | | 1 | | | - | | | - | | | 1 | | |
| **PPMS** | 5 | | | 1 | | | - | | | 6 | | | 1 | | |
| **QC exclusions** | 5 | | | 1 | | | 3 | | | 2 | | | 4 | | |

dMRI = diffusion MRI; rs-fMRI = resting-state functional MRI; TR = repetition time; TE = echo time; TI = inversion time; FA = flip angle; EPI = echo-planar imaging; PEPOLAR = Phase-encoding polarity; CIS = clinically isolated syndrome; HC = healthy controls; PPMS = primary-progressive multiple sclerosis; QC = quality control; RRMS = relapsing-remitting multiple sclerosis; SPMS = secondary-progressive multiple sclerosis; QC = quality control.

**Supplementary Table 2. Results of the between-group comparison in terms of multiplex regional coreness.** Regions for which a significant difference emerged at the PwMS vs HC comparison are shown, along with corresponding effect sizes (Hedges' g) and FDR-adjusted *p* values. The nomenclature of cortical areas follows the 7-network Schaefer-100 parcellation.[1]

| ROI name | Hedges' g | FDR-adjusted *p* |
|---|---|---|
| LH_Thal | 0.91 | 0.003 |
| LH_Cau | 0.39 | 0.003 |
| LH_Put | 0.38 | 0.003 |
| LH_Pall | 0.24 | 0.003 |
| RH_Thal | 0.94 | 0.003 |
| RH_Cau | 0.37 | 0.003 |
| RH_Put | 0.30 | 0.003 |
| LH_Cont_Cing_1 | -0.19 | 0.005 |
| RH_Default_PFCdPFCm_1 | -0.21 | 0.005 |
| RH_Pall | 0.21 | 0.01 |
| RH_Amyg | 0.18 | 0.01 |
| RH_Hipp | 0.19 | 0.02 |
| LH_Default_Temp_2 | -0.17 | 0.04 |
| LH_Amyg | 0.17 | 0.05 |

**Supplementary Figure 1. Average connectivity matrices in the healthy controls group.** Matrices of (**A**) morphological covariance, (**B**) structural connectivity, and (**C**) functional connectivity averaged over all healthy subjects. Matrices are ordered according to 7 canonical resting-state networks,[1] plus a network of subcortical gray matter regions.

VIS = visual network; SM = somatomotor network; DAN = dorsal attention network; VAN = ventral attention network; L = limbic network; CONT = control network; DMN = default mode network; SUBC = subcortical network.

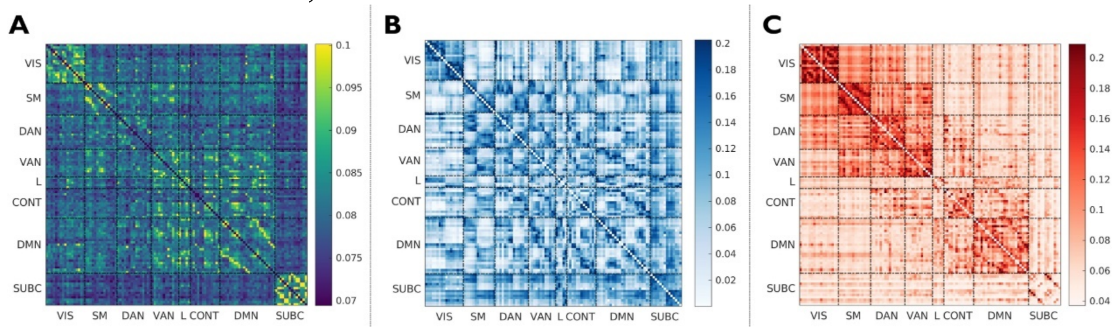

**Supplementary Figure 2. Average single-layer coreness in the healthy controls group.** Color-coded (*teal* to *red*) maps of coreness percentile ranks in the (**A**) morphological covariance, (**C**) structural connectivity, and (**E**) functional connectivity domains, superimposed on surface renderings of the cortex and subcortical structures. Images were obtained with the ENIGMA toolbox.[2] (**B-D-F**) Highest 10% coreness nodes and corresponding absolute values are shown for the different layers. Nomenclature of cortical areas follows the 7-network Schaefer-100 parcellation.[1]

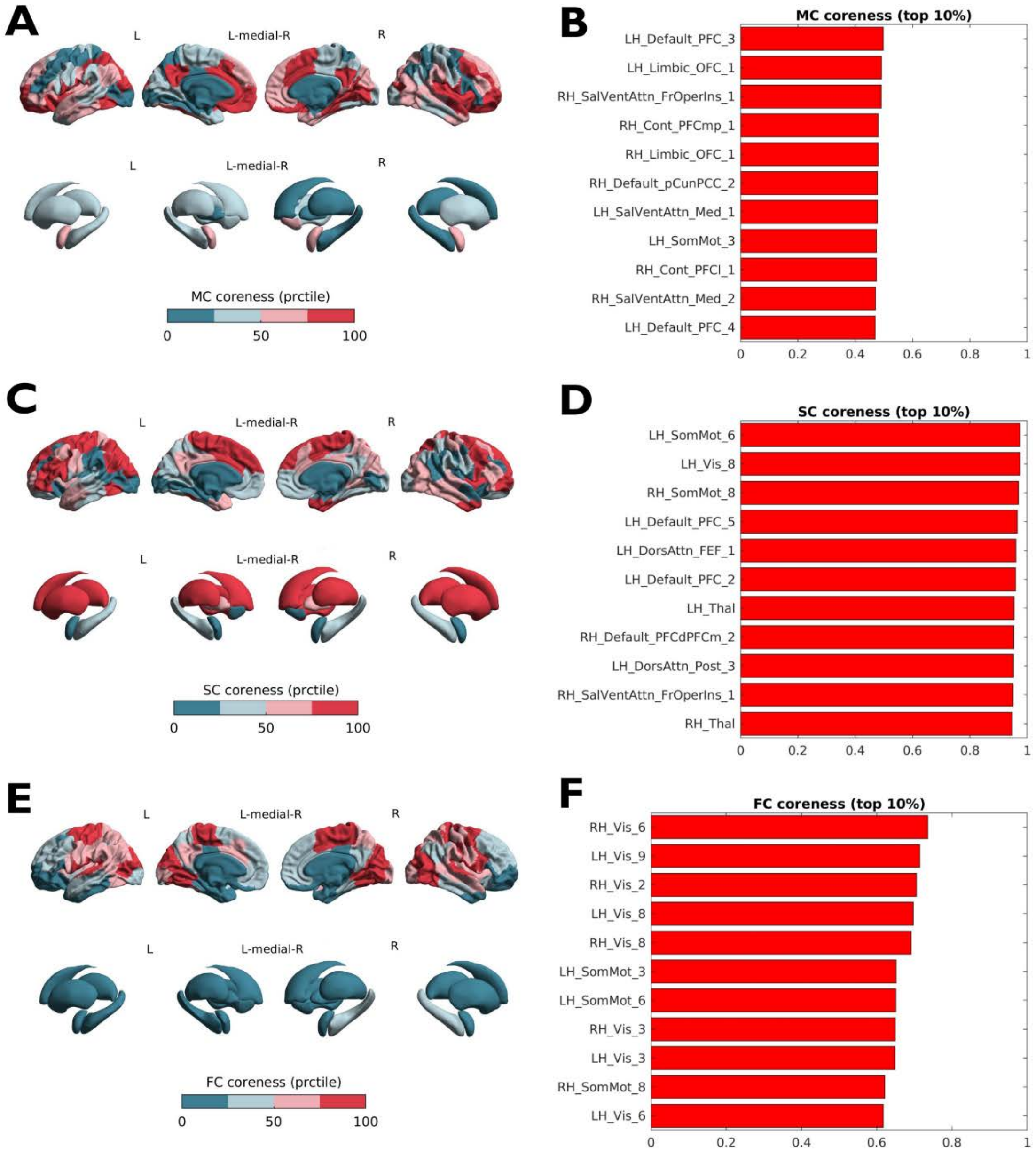
